# Social Determinants Associated with COVID-19 Mortality in the United States

**DOI:** 10.1101/2020.08.28.20183848

**Authors:** Shayom Debopadhaya, Ariella D. Sprague, Hongxi Mou, Tiburon L. Benavides, Sarah M. Ahn, Cole A. Reschke, John S. Erickson, Kristin P. Bennett

## Abstract

This study examines social determinants associated with disparities in COVID-19 mortality rates in the United States. Using county-level data, 42 negative binomial mixed models were used to evaluate the impact of social determinants on COVID-19 outcome. First, to identify proper controls, the effect of 24 high-risk factors on COVID-19 mortality rate was quantified. Then, the high-risk terms found to be significant were controlled for in an association study between 41 social determinants and COVID-19 mortality rates. The results describe that ethnic minorities, immigrants, socioeconomic inequalities, and early exposure to COVID-19 are associated with increased COVID-19 mortality, while the prevalence of asthma, suicide, and excessive drinking is associated with decreased mortality. Overall, we recognize that social inequality places disadvantaged groups at risk, which must be addressed through future policies and programs. Additionally, we reveal possible relationships between lung disease, mental health, and COVID-19 that need to be explored on a clinical level.

## Introduction

The widespread prevalence of COVID-19 has forced the pandemic to be the center of state and national policy in the United States (US). As of July 5, 2020 (the date of this study), the CDC has documented over 3 million infections and over 130,000 COVID-19 deaths in the US [1]. These deaths are not uniformly distributed, and several counties are experiencing higher-than-average death rates [2]. It is hypothesized that social determinants have contributed to these disparities in COVID-19 mortality [3], which we investigate. Social determinants, such as a county’s access to healthcare, rates of education, indicators of health, and economic status have greatly impacted other diseases [4], so these factors may play a similar role in COVID-19.

A growing amount of literature highlights that social determinants of COVID-19 place socioeconomically disadvantaged populations at increased risk. Abrams and Szefler reviews emerging literature to discuss the stark inequality of COVID-19 infection rates and outcomes among several groups [3]. This work recognizes that social determinants such as housing problems, race, smoking, nutrition, overcrowding, poverty, and comorbidities may compromise an individual to COVID-19. Ahmed et al. is another among the many to describe the disproportionate effects of COVID-19 among the socioeconomically disadvantaged, highlighting access to healthcare as one of the many determinants that are potentially critical in COVID-19 mortality [5]. Fielding-Miller et al. studies a few select social determinants to find that dense population in urban counties and non-English speakers, farm workers, and impoverish groups in non-urban counties are at increased risk [6]. Overall, the literature emphasizes that quantifying the social determinants of COVID-19 is a crucial step in addressing the existing health inequalities. Yet, there is a lack of well-controlled, diverse screening of the social determinants of COVID-19.

To address the gap in the current literature and leverage readily available data, this study identifies which social determinants are associated with a significant changes in COVID-19 mortality rate at the county level. Uniquely, we account for a comprehensive list of comorbidities and the impact of differing state policies. The study consists of an initial ecological overview to establish which high-risk factors from literature are significant. Then, using the significant high-risk factors as controls, 41 social determinants are evaluated to find those that affect COVID-19 mortality. Both the initial overview to identify controls and the association study utilize negative binomial mixed models to analyze county level data (n=3093). Results are statistically corrected for possible false discoveries. Further sensitivity analysis with different model variants and time-series analysis are validated.

## Methods

All methods and results are reproducible. The full R code and a supplementary document with additional details can be found on the Rensselaer IDEA Github.^1^

### Identifying High-Risk Controls

A robust set of controls needed to be established before screening for social determinants [7]. To find the most important risk factors, we considered the guidelines from the Center of Disease Control (CDC). Using extensive metaanalysis, the CDC recognized that those suffering from several disease groups and ethnic minorities as being at higher risk for developing severe illness and death due to COVID-19 [8]. Our analysis accounts for these well-documented risk factors, using available data for the racial distribution in the US, as well as data for several of the highest risk comorbidities, such as the prevalence of cardiovascular death, obesity, diabetes, COPD, lung cancer, and asthma. Additionally, temporal variables such as days since first infection were included to adjust for disease progression in each state, as this would greatly affect mortality rates of any region. Income and education were used as initial controls for socioeconomic status [5]. A breakdown of population density by quartile was used as a proxy to categorize the urban or rural identity of a county. Variables were scaled by subtracting the mean from each data point and dividing by the standard deviation.

Using county-level data for the entire U.S. updated as of July 5, 2020, a negative binomial mixed model was used to find the significant high-risk factors. The modeling method and observed and explanatory variables were a generalization of those used in a prior study on the impact of air pollution on COVID-19 mortality [9]. COVID-19 death rate was used as the observed variable. The following explanatory variables were used: percent of county that is African American, percent of county that is Hispanic, percent of county that is Native American, percent of county that is Asian, percent of county that is White, median household income of a county, percent of county with less than a high-school education, percent of county that is above the age of 65, population density in quartiles, days since first infection, days since social isolation, days since mask required, days since reopening, days since reclosure, and available hospital beds per county population. Additionally, to account for potential COVID-19 comorbidities, the model used percent of county that is obese, percent of county that has diabetes, cardiovascular death rate, percent of county with lung cancer, percent of county with COPD, percent of adults in county with asthma. An offset to scale for population and a term to account for random variance at the state level was also included [9, 10].

The extracted output from this model is the the ratio of change in COVID-19 death rate per unit increase in each explanatory variable. We refer to this output as Mortality Rate Ratio (MRR) [9]. In this initial analysis, these explanatory variables reveal the important baseline effects that must be controlled for when screening for social determinants.

### Screening for Social Determinants

The statistically significant terms from the methods above were used as controls for the social determinant screening. This study design identifies unique social determinants that are distinguishable from the already recognized high-risk factors.

For the social determinant screening, negative binomial mixed models were used in the association analysis to individually test 41 variables that represented a wide range of specific social determinants. This set included metrics of a county’s mental health, physical health, rates of disease, economic status, housing burdens, demographics, and multiple death rates. These determinants are mainly sourced from County Health Rankings. The model produces an MRR for each determinant. A complete list of the explanatory variables and sources can be found in the supplementary document.

### Statistical Testing

Each explanatory variable in the high-risk model and each variable in the 41 association models was tested for statistical significance using a Wald test [11]. The significant p-values are shown with their corresponding coefficients in Tables 1 and 2. The Benjamini-Hochberg Procedure [12] was used to adjust p-values for Multiple Hypothesis Testing in the social determinant screening, which is reflected in the results of Table 2. Our models showed robustness in this process, as the false discovery rate was acceptably below 0.05 at 0.0217.

**Table 1:** Statistically Significant Results From The High Risk Model. The coefficients (left column) is the name of the explanatory variable. The MRR (center column) is the change in COVID-19 mortality rate per unit increase of explanatory variable. The p-value (right column) is the level of significance returned for each term in the model.

| Coefficients | Mortality Rate Ratio (percent increase in COVID-19 mortality rate per standard deviation increase of coefficient) | p-value |
| --- | --- | --- |
| Days Since First Infection | 124.94 | <2.00e-16 |
| Percent of County with Lung Cancer | 40.87 | 5.11e-03 |
| Percent African American in County | 36.10 | 2.72e-02 |
| Days Since Mask Required | 30.41 | 8.36e-03 |
| Median Household Income | 22.00 | 8.33e-06 |
| Percent Hispanic in County | 21.53 | 1.30e-04 |
| Percent Native American in County | 16.34 | 2.12e-02 |
| Percent with less than High-School Education | 14.13 | 1.46e-03 |
| Percent Asian in County | 8.31 | 3.53e-02 |
| Percent of County with Adult Asthma | -20.06 | 2.01e-02 |

**Table 2:**
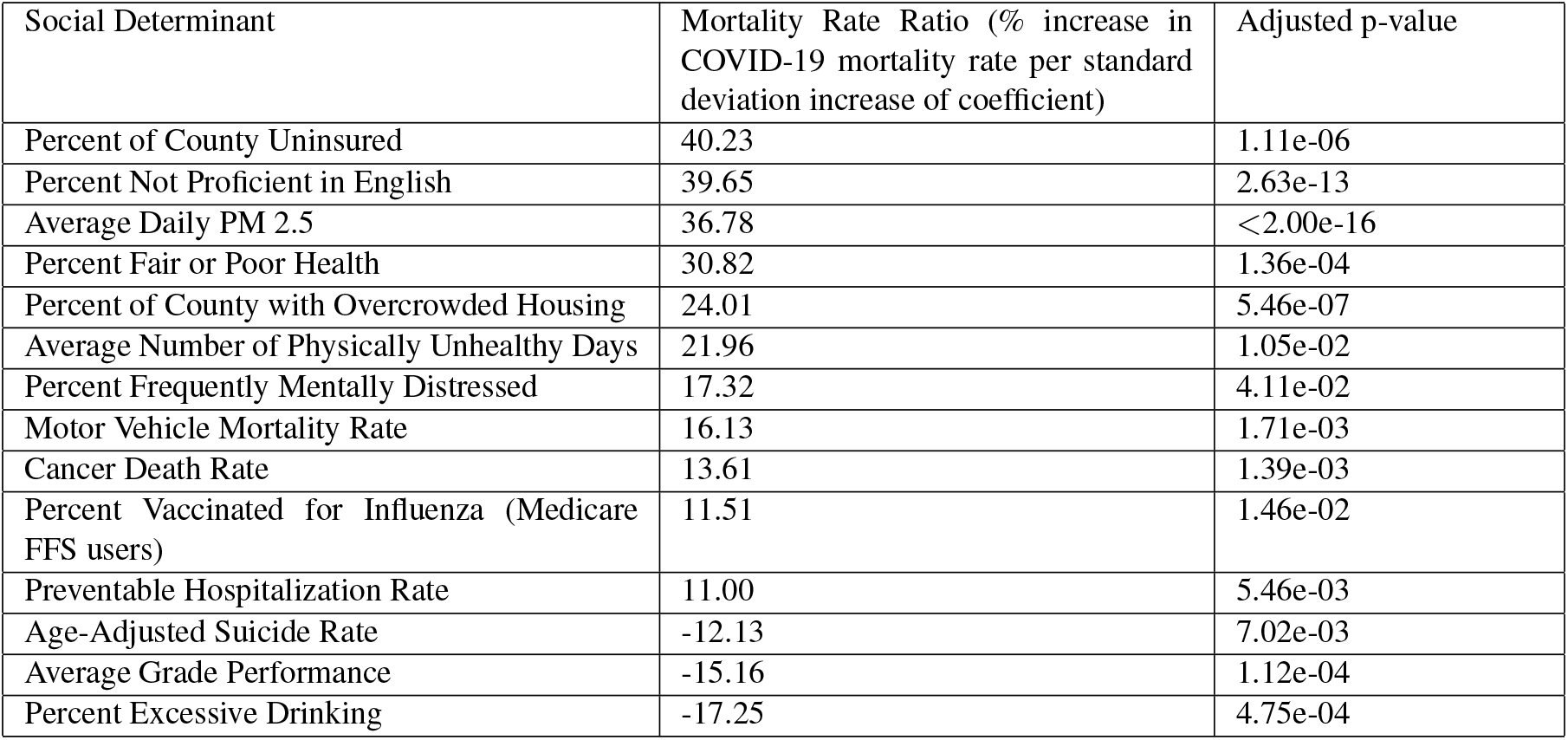
Statistically Significant Results From The Association Analysis. The social determinant (left column) is the name of the social determinant tested in the model. The MRR (center column) is the change in COVID-19 death rate per unit increase of social determinant. The p-value (right column) is the level of significance returned for each term in the model, adjusted with the Benjamini-Hochberg Procedure.

| Social Determinant | Mortality Rate Ratio (% increase in COVID-19 mortality rate per standard deviation increase of coefficient) | Adjusted p-value |
| --- | --- | --- |
| Percent of County Uninsured | 40.23 | 1.11e-06 |
| Percent Not Proficient in English | 39.65 | 2.63e-13 |
| Average Daily PM 2.5 | 36.78 | <2.00e-16 |
| Percent Fair or Poor Health | 30.82 | 1.36e-04 |
| Percent of County with Overcrowded Housing | 24.01 | 5.46e-07 |
| Average Number of Physically Unhealthy Days | 21.96 | 1.05e-02 |
| Percent Frequently Mentally Distressed | 17.32 | 4.11e-02 |
| Motor Vehicle Mortality Rate | 16.13 | 1.71e-03 |
| Cancer Death Rate | 13.61 | 1.39e-03 |
| Percent Vaccinated for Influenza (Medicare FFS users) | 11.51 | 1.46e-02 |
| Preventable Hospitalization Rate | 11.00 | 5.46e-03 |
| Age-Adjusted Suicide Rate | -12.13 | 7.02e-03 |
| Average Grade Performance | -15.16 | 1.12e-04 |
| Percent Excessive Drinking | -17.25 | 4.75e-04 |

To test performance and fit, the procedure from Wu et al. was followed [9]. The negative binomial mixed model for the high-risk factors was compared to zero inflated, fixed negative binomial, and spatial correlation variants. These models were compared to the negative binomial mixed model using AIC and BIC. The main high-risk model was also analyzed at 16 different time points for robustness. Additionally, variants of the model that excluded New York city data and excluded counties with less than 10 cases were analyzed for robustness.

Statistical variations produced no significantly different results. Model was robust to time-series analysis. Any changes in results due to different sample stratification can be found in the supplementary document.

All analysis was conducted in R [13] using the lme4 package. Notebooks to reproduce all results can be found on the Rensselaer IDEA Github.

This social determinants analysis is included in COVIDMINDER^2^, an online tool for exploring disparities in the COVID-19 epidemic. COVIDMINDER is run by the Rensselaer Institute for Data Exploration and Applications at Rensselaer Polytechnic Institute.

### Data Source

A conglomerate of data was formed from the CDC, County Health Rankings [14], and JHU [15]. Detailed information about the data sources is included in the online supplementary document.

## Results

### High-Risk Control Model

The high-risk model reveals several statistically significant factors. These results are presented in Table 1.

### Social Determinant Screening

The significant variables mentioned above were included as controls in the social determinant screening. Lung disease was not included, as we identified an anomaly in our results, which we address in the discussion section. For completeness of variables, data from all races and all quartiles of population density were included. Hence, the Social Determinant Screening Procedure had the following design:

COVID-19 Mortality Rate – percent African American in the county + percent Hispanic in the county + percent of Native Americans in the county + percent Asian in the county + percent White in the county + population density in quartiles + median household income + percent without high school education + beds/population + days since first infection + days since mask required + offset(population) + (term for random variation by state) + (SOCIAL DETERMINANT)

41 iterations of this model were analyzed, each using a different social determinant. The p-values were adjusted by Benjamini-Hochberg to account for multiple hypothesis testing. The MRR and adjusted p-values of the statistically significant social determinants from their respective models are reported in Table 2. Note that a full list of the social determinants can be found in the supplementary document.

## Discussion

This study conducts an analysis of social determinants and comorbidities at the national scale with a wide variety of controls and county level data. We reveal the high-risk factors and the specific social determinants of a county that affect the mortality rate of COVID-19.

### Identified High-Risk Factors

Our results suggest that ethnic minorities have an increased risk to COVID-19. Counties experience 36.11%, 21.53%, 16.34%, and 8.31% increases in COVID-19 mortality per standard deviation increase in African American, Hispanic, Native American, and Asian representation, respectively. Historically, these ethnic minorities have disadvantaged access to healthcare, occupational opportunities, higher chronic stress, lower access to quality education, and more housing problems [16]. We conclude these racial inequalities have a role in the increased COVID-19 mortality rate.

One standard deviation increase in percent of county population without high school education is associated with a 14.13% increase in COVID-19 MRR. Education is a well-studied determinant of adolescent health [17], and lower high school graduation has been specifically linked to poorer health outcomes in the past [18]. We observe the same phenomenon in the context of COVID-19. This finding is further substantiated by the fact that higher average grade performance is associated with a -15.16% change in COVID-19 MRR, as seen in Table 2. Average grade performance is defined as the outcomes of 3rd grade reading scores compared to the national average [19]. This is a metric for available educational opportunities [20], a predictor of future academic performance [21], and a predictor of future health outcomes [22, 23, 24]. Our results suggest that educational disparity has a significant role in mortality rate outcomes in the COVID-19 pandemic.

The model reports a 40.87% increase in MRR per standard deviation increase in lung cancer prevalence. While the CDC already recognized lung cancer as a high-risk factor [8], this data suggests that lung cancer may be of the most serious comorbidities for COVID-19. Clinical data supports this phenomenon, and one study even describes that 52.3% of COVID-19 patients with lung cancer died, compared to the 192 deaths out of the 1878 total COVID-19 patients [25].

The temporal variables of days since first infection and days since mask required have MRRs of 124.94% and 30.41%, respectively. These represent the disease progression and adjust the results for counties that experienced high infection rates at different times. We observe that an early incidence of COVID-19 in a county is associated with the greatest increase in COVID-19 mortality. It is possible that as the pandemic progressed, the improved clinical experience of treating COVID-19, appropriate resources, and better compliance with social distancing may be responsible for this finding. It is also likely that regions with higher numbers of COVID-19 cases and deaths also required masks earlier, which is why the corresponding MRR is so high. Rather than days since masks required as a causal factor of COVID-19 mortality, we recognize it as a covariate.

### Additional Findings About High-Risk Factors

One standard deviation increase in county median income is associated with a 22.00% increase in COVID-19 mortality rate per standard deviation. In a traditional view of social determinants, higher income is associated with lower rates of disease [26]. One explanation for our finding between income and COVID-19 is the role of urban counties, as income is higher in urban counties [27]. Our results may provide evidence of a heightened urban risk to COVID-19.

Surprisingly, one standard deviation increase in adult asthma prevalence is associated with a 20.06% decrease in mortality rate (negative MRR), whereas the CDC identified asthma as a possible comorbidity [28]. The observed protective role of asthma may be due to an interaction between inhaled corticosteroids (ICS) and COVID-19 [29, 30]. Preliminary literature is currently mixed [31, 32] and calls for epidemiological study with detailed information on comorbidities [30]. It is hypothesized that a decrease in angiotensin-converting enzyme 2 (ACE-2), a receptor for SARS-CoV-2, may be associated with the use of inhaled corticosteroids, a possible treatment for asthma [29]. Our study uses a robust set of comorbidities and confounds, so it is suspected that the relationship we find between adult asthma prevalence and COVID-19 mortality rate may have significance at the clinical level. However, since there is a current lack of clinical work that establishes a thorough basis for this relationship, we excluded lung disease from the controls in the association study to avoid statistical artifacts in our results.

The other factors we tested are statistically insignificant in the high-risk model. Diabetes, age above 65, obesity, and COPD have all been documented in literature to have unfavorable interactions with COVID-19 [8], but we find that they lack significance in aggregate data. Including a large number of terms in the model may have diminished the effect of these factors. It is also possible that these factors may be regionally significant, and further analysis is needed at the sub-national level.

### Social Determinant Screening

The following is a detailed discussion of the significant social determinants from the 41 factor association study, which are presented in Table 2.

#### Poor Healthcare

One standard deviation increase in preventable hospitalization rate and percent of uninsured population is associated with 11.00% and 40.23% increases in MRR, respectively. These determinants are direct indicators of quality of healthcare. Overuse of hospitals typically indicates that outpatient care is inadequate or that there is limited available primary care [33]. Counties that were ill-equipped for their population pre-pandemic may continue to be inadequate to their populations during a pandemic, leading to more COVID-19 deaths. A high percent of uninsured individuals also indicates poor access to health care. In general, those without access to health insurance will not be given thorough preventative care, often resulting in undiagnosed health problems or severe illness [34], which may result in more COVID-19 deaths.

One standard deviation increase in percent of county with frequent mental distress, average number of physically unhealthy days in the county, and percent of county population with poor or fair health is associated with a 17.32%, 21.96%, 30.82% increase in COVID-19 MRR, respectively. Our findings suggest that counties with poor health outcomes before the pandemic have poor COVID-19 outcomes. Unhealthy counties may have a high prevalence of common underlying health conditions, which places them at increased risk to COVID-19, as recognized by the CDC[35]. Poor access to healthcare also has a likely role in the increased COVID-19 mortality rate in these counties, as poor mental and physical health has been linked to socioeconomic disadvantage [36, 37].

One standard deviation increase in cancer death rate is associated with a 13.61% increase in COVID-19 mortality rate. Higher cancer death rate can result both from increased incidence and unfavorable outcomes due to poor access to healthcare. High cancer mortality rates indicate that a community has insufficient prevention, early diagnosis, and treatment [38]. Low income communities are notably at a disadvantage for cancer treatment due to a lack of resources, as wealth disparities are noted as the most common cause of health disparities [39].

#### Immigrant Communities

One standard deviation increase in percent of individuals not proficient in English is associated with a 39.65% increase in COVID-19 mortality. As of 2013, the percentage of Limited English Proficient (LEP) people who were foreign born was 81.3% [40]. These results suggest that immigrant populations are at higher risk from COVID-19, which is supported by previous work [6]. Historically, immigrants suffer from poverty, and many immigrants are barred from receiving healthcare programs such as Medicare in their first 5 years of living in the US. Additionally, undocumented immigrants are not eligible for any public programs. It is estimated that around 45% of documented immigrants and 65% of undocumented immigrants do not receive healthcare. This leaves the population much more vulnerable. LEP immigrants also receive poorer quality of care and understanding of their medical condition due to language barriers in treatment. Overall, immigrant populations are at a much higher risk for severe outcomes due to lower access and poorer quality of care.

#### Poverty & Severe Housing Problems

One standard deviation increase in overcrowding, a measure of of poor socioeconomic status [41], is associated with a 24.01% increase in COVID-19 MRR. This data is defined as the ratio of residences with more than 1 person per room in living circumstances to the total number of housing units in a county. Overcrowding is a metric of severe housing problems, previously associated with poor mental health outcomes [42], tuberculosis [42], and several other diseases [43]. Beyond signifying poor socioeconomic status, there is also physical risk with any infectious disease from overcrowding due to close proximity of potential carriers [44]. These attributes of poverty and housing problems are associated with a significant increase of COVID-19 mortality across the nation.

#### Air Pollution

One standard deviation increase in average air pollution is associated with a 36.78% increase in MRR. The significance of air quality (PM2.5) in the social determinant screening supports Wu et al.’s results, indicating that counties with long-term air pollution are more likely to fall severely ill [9].

#### Vaccination Rates in Medicare FFS

One standard deviation increase in influenza vaccination rates among Medicare Fee For Service (FFS) users is associated with a 11.51% increase in COVID-19 MRR. One explanation for this result is that elderly groups in community care and those with pre-existing conditions like cardiovascular disease vaccinate more frequently. 83.1% of Medicare FFS users are above the age of 65 [45], and flu vaccinations are more common in individuals above 65 years old (72.3%) compared to those between 18 and 64 years old (30.7%) [46]. Flu vaccination rates are also higher in patients with comorbidities like cardiovascular disease in comparison to patients without cardiovascular disease [47]. As elderly populations and groups with comorbidities are more vunerable COVID-19 [8], the observed relationship between between vaccination rates and COVID-19 may represent the underlying relationship between at-risk groups and COVID-19. However, as there are severe geographical, racial, and socioeconomic disparities in influenza vaccination rates [48], future analysis of vaccination and COVID-19 mortality is needed to reveal more about this complex relationship.

#### Motor Vehicle Mortality Rate

One standard deviation increase in motor vehicle mortality rate is associated with a 16.13% increase in COVID-19 MRR. Increased motor vehicle mortality rate has been associated with counties with high uninsurance and high concentrations of ethnic minorities[49]. As listed above, we find these groups to have an increased risk to COVID-19. However, increased motor vehicle mortality rate is also associated with rural counties and populations under the age of 18 [49], which we did not find to be significantly associated with increased COVID-19. It is possible that the relationship between motor vehicle mortality rate and COVID-19 has factors beyond the scope of this study, so future work is needed to fully understand this relationship.

#### Suicide and Excessive Drinking

We observe a statistically significant, negative relationship between age-adjusted suicide rate and excessive drinking with COVID-19 mortality rate (−12.13 MRR and -17.25 MRR, respectively). This suggests a complex relationship between the pandemic and the recent epidemic of “deaths of despair”: deaths from suicide, overdose, alcoholism, and self-harm. Alcohol is a coping strategy for mass stress and isolation, and consumption increased greatly after the pandemic began [50]. Suicide rates are higher in less urban areas and lower in more urban areas while the reverse is true for COVID-19 mortality rates [51]. While supporting effective social distancing, social isolation is a major risk factor for suicide [52]. The inverse relationship of pre-COVID-19 suicide and alcohol consumption with COVID-19 deaths are likely only temporary as suicide rates and alcohol abuse are likely to increase during the pandemic [53]. Further work is needed to fully understand the relationships that we observe.

## Conclusion

Overall, we identify that counties with more ethnic minorities, more immigrant populations, lower high-school graduation rates, higher lung cancer prevalence, worse healthcare, higher poverty, higher motor vehicle mortality, earlier exposure to COVID-19, higher vaccination rates among Medicare FFS users, and those in urban environments have an increased risk of COVID-19 mortality. Like other diseases [4], the severity of COVID-19 is greatly worsened by socioeconomic inequality. Interventions and policies are needed to protect these disadvantaged populations during the ongoing pandemic. COVID-19 also serves as evidence that long-term measures to address growing health inequalities [37] are necessary.

Care must be taken in contextualizing these results due to inherent limitations of county-level ecological studies. While relationships observed in aggregate data cannot be translated from the county level to individuals from an ecological study alone, these studies identify useful trends and hypotheses for future analysis at the individual level [54]. Accounting for possible confounds is also essential in ecological studies, which we achieve by including several controls in every model. Other limitations of the study stem from the availability and accuracy of the data. Literature suggests that both the reported counts of infection and the reported COVID-19-related deaths have inconsistencies [55, 56], and the lack of proper testing has also been highlighted [57].

As more detailed information becomes available, it would prove useful for future study to verify our results using clinical data. Further investigation of the relationship between COVID-19 and adult asthma may be especially valuable, as there is currently varying evidence. Additionally, the relationships between COVID-19, motor vehicle mortality rate, vaccination rate, suicide, and alcohol consumption need to be elucidated in more detail. These analyses may reveal further interactions between social determinants and COVID-19 mortality.

## Data Availability

A publicly-accessible repository containing all data used in this work has been available and is cited in the paper. All data used in this work was obtained via public sources.

https://github.com/TheRensselaerIDEA/COVIDMINDER/tree/master/social-determinants-paper

## Acknowledgements

This study was supported by the Rensselaer Institute for Data Exploration and Applications, the Data INCITE Lab, and a grant from the United Health Foundation.

## Footnotes

1 https://github.com/TheRensselaerIDEA/COVIDMINDER/tree/master/social-determinants-paper

2 https://covidminder.idea.rpi.edu/ under the NATIONAL REPORT CARD tab

